# CONVEX VERSUS LINEAR TRANSDUCER IN COVID-19 LUNG ULTRASOUND: A PROSPECTIVE COMPARATIVE COHORT

**DOI:** 10.1101/2025.01.31.25321454

**Authors:** Jaime Gil-Rodríguez, José-Antonio Peregrina-Rivas, Pablo Aranda-Laserna, Michel Martos-Ruíz, Miguel Ángel Montero-Alonso, Alberto Benavente-Fernández, Juan Melchor, Emilio Guirao-Arrabal, José Hernández Quero

**Affiliations:** Internal Medicine Unit. San Cecilio University Hospital. 18016, Granada. Spain; Infectious Diseases Unit. San Cecilio University Hospital. 18016, Granada. Spain; Department of Statistics and Operations Research. University of Granada. 18016, Granada. Spain; Biomechanics Group (TEC-12). Instituto de Investigación Biosanitaria (IBS), 18012 Granada, Spain Research Unit “Modelling Nature” (MNat). University of Granada, 18011 Granada, Spain

**Keywords:** SARS-CoV-2, COVID-19, lung, ultrasound, LUS

## Abstract

**Background and purpose:** It is currently unknown whether ultrasound transducer differences influence COVID-19 pneumonia images. Our aim was to describe and compare the results obtained using convex and linear transducers for image acquisition in COVID-19 lung ultrasound.

**Methods:** This is a single centre prospective cohort study, comparing convex and linear probes in 16 adult patients with SARS-CoV-2 pneumonia. We analysed 12 lung-fields and described the presence of pleural thickening, pleural irregularity, B-lines, >3 B-lines, coalescing B-lines, subpleural consolidation, pulmonary consolidation and pleural effusion in each of these fields and with both transducers. In addition, the Lung Ultrasound Score (LUS) was calculated for every patient using the two transducers.

**Results:** The mean LUS difference between both transducers was statistically significant in the right lower lateral, upper and lower posterior quadrants; as well as in the left lower anterior and upper posterior quadrants. Furthermore, mean total LUS difference was also statistically significant (15.81 convex vs. 10.81 linear; p = 0.001). The most frequently described findings with the convex transducer compared to the linear transducer were pleural irregularity, pleural thickening, >3 B-lines, coalescing B-lines and subpleural consolidation. On the opposite side, pleural effusion was more frequently described with the linear probe.

**Conclusions:** Lung ultrasound in COVID-19 pneumonia is a transducer-dependent technique. This difference is more pronounced in posterior quadrants and in the cardiac area. Although both linear and convex transducers are suitable for the evaluation of COVID-19 pneumonia, the convex probe provides more information than the linear probe if only one of the two is accessible.

## 1. Background

As of March 3rd, 2023, more than 676 million people have been infected by SARS-CoV-2 with a total dead 6.8 million (over 1% of the total infections) [1], The study of pneumonia caused by SARS-CoV-2 (COVID-19) is highly relevant, as it is characterized by strong variability, which increases the difficulty of diagnosis [2]. Hence, several tests and prognostic scores have been validated to facilitate COVID-19 management. Among these scores, those provided by thoracic imaging techniques have been considered essential in the diagnosis of COVID-19 pneumonia [3].

Lung ultrasound has been demonstrated to be a rapid, inexpensive, reproducible, and more sensitive diagnostic procedure for COVID-19 pneumonia than other thoracic imaging tests [4]. Evidence supporting the use of lung ultrasound in COVID-19 evaluation is increasing, and it has become an acknowledged alternative to chest CT scans for both diagnosis and for establishing the severity of the disease [3, 4, 5, 6, 7].

Many severity scores have been developed, most of them considering clinical and analytical parameters. However, only some of them include lung ultrasound as a predictor of worse outcomes and of mortality [8]. Based on the use of B lines to discern the severity of lung inflation, different scores have been established in recent years to calculate a quantitative and exhaustive indicator as a prognostic marker [9, 10, 11, 12, 13]. Different proposals have emerged such as B-line number, LUS, COWS, and artificial intelligence-based methods [14, 15, 4, 16, 17, 18, 19]. Lung ultrasound scores, such as those proposed by Soldati et al. based on Rouby and Soummer’s previous studies [20, 21, 22], are the most widely used for COVID-19 pneumonia evaluation. This score is still the standard used in clinical practice, and the one recommended in some review articles on lung ultrasound in COVID-19 [23]. It has also been independently validated in subsequent studies [24, 25].

However, no determined protocol for lung ultrasound use has been established yet [26, 27]. Regarding transducers, there are no comparisons of the advantages between them, to establish one as a reference. Moreover, ultrasound devices are not always equipped with convex and linear probes, and oftentimes there is not enough time for both probes to categorize the results effectively. Although the mode of transmission is analogous [28, 29], the recommendations are indistinct [30].

As a result, it would be interesting to know the advantages of one probe over the other, to optimize the time spent on lung ultrasound as much as possible. This has a special interest in a pandemic context, where all kinds of resources must be optimized. Although this problem is rarely studied in the field of adult lung ultrasound, it is well-known in the field of neonates [31]. So, the present study aims to describe the results obtained when using a convex or linear transducer and to establish the benefits of one probe over the other in COVID-19 pneumonia evaluation.

## 2. Methods

### 2.1. Source of data

A single-center prospective cohort study to compare the convex and linear probes. Our study protocol was approved by the regional ethics committee (0259-N-21 code) and each participant provided informed consent to participate in the study. We wrote this article by the TRIPOD statement for risk prediction models [32].

### 2.2. Participants

This study was conducted at a secondary hospital. Participants were consecutive adult patients (aged 18 years or older) admitted to the COVID-19 inpatient wards in the first 24 hours. All infections were confirmed by real-time reverse transcription. polymerase chain reaction (RT-PCR) SARS-CoV-2, from 5th May to 13th July 2021. As exclusion criteria, patients with suspected or confirmed bacterial pulmonary superinfection, those unable to cooperate with the examination, and those who voluntarily refused to participate, were missed out on the study. An informed consent and study summary form were given to all patients, who signed it with the possibility of revocation.

### 2.3. Patient characteristics and clinical outcomes

In the study, the following variables were collected from each patient: registration data, anthropometric variables, symptoms, comorbidities, vital signs, analytical data, and ultrasound lung involvement. Analytical data were requested at first hospital contact, whereas clinical variables and missing data were collected by the researchers on the COVID-19 transitional hospitalization ward at the time of evaluation. The outcomes measured were non-invasive mechanical ventilation or high-flow nasal oxygen therapy (NIMV), invasive mechanical ventilation (IMV), intensive care unit (ICU) admission, and 28-day mortality. We also elaborated on the combined variable poor outcome (NIMV or IMV or ICU admission or 28-day mortality). The primary variable of this study was the combined variable “poor outcome”, and the secondary variable “28-days mortality”.

### 2.4. Image acquisition

The ultrasound scanner was PHILIPS®SPARQ (Koninklijke Philips N.V). We established the predefined lung preset, using both a curved array transducer of 6-2 MHz and a linear array transducer of 12-4 MHz. Four Internal Medicine physicians, members of the Clinical Care Ultrasound Group (recognized with the excellent certificate of the Spanish Society of Internal Medicine and trained in COVID-19 pneumonia) performed the examinations, divided into two groups of two operators respectively. Each group carried out half of their scans with the convex transducer and the other half with the linear one, and each operator performed the same number of acquisitions with both probes. This way, potential bias associated with the inter-individual variability was equally distributed. The images acquired from each probe were obtained consecutively in the same patient, blinding the report of the exploration results or consultation of the patient’s history. We used the 12-lung field protocol, as the searching strategy with the most efficient balance between diagnostic accuracy and acquisition time [33]. Each hemithorax was divided into anterior, lateral, and posterior fields by the anterior and posterior axillary lines. Afterward, each space was divided into upper and lower fields, with a total of 6 fields for each hemithorax. Ultrasound findings were recorded using a template common to all investigators. This included, for each of the 12 fields analyzed, the presence or absence of the following: pleural thickening, pleural irregularity, B-lines, ¿ 3 B-lines, coalescing B-lines, subpleural consolidation, pulmonary consolidation, and pleural effusion. We also established the Lung Ultrasound Score (LUS) in each of these fields, as well as the total score summed over all of them [20] (Figure 1).

**Figure 1:**
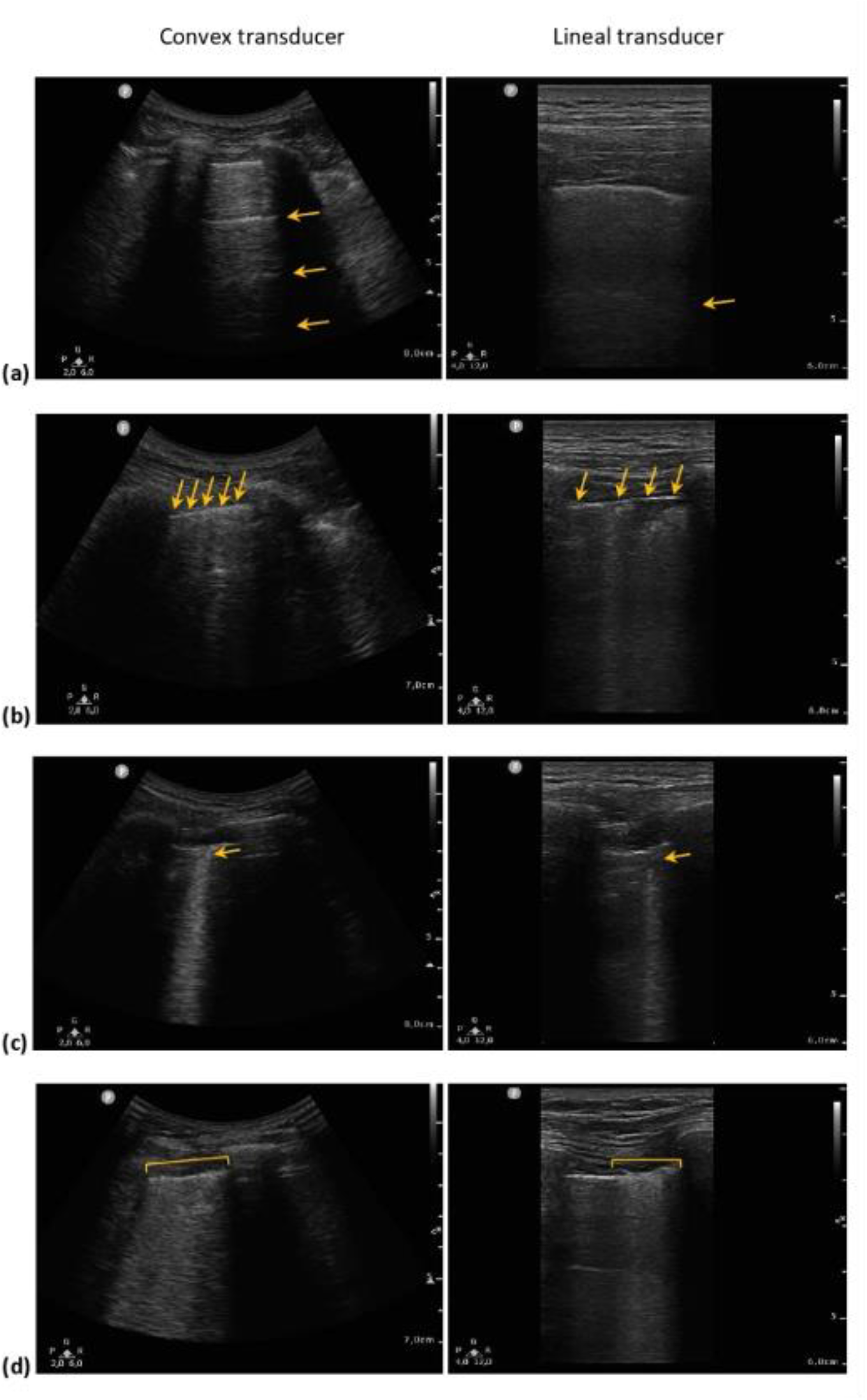
Comparison between LUS obtained by convex (column 1) and linear (column 2) probes respectively in the same LUS field and patient: (a) 0 points [normal pattern of A-lines]; (b) 1 point [> 3 B-lines]; (c) 2 points [subpleural consolidation]; (d) 3 points [pulmonary or “with lung” consolidation].

High-frequency probes as linear transducers provide higher resolution, but a shallower depth of field. Low-frequency transducers the case of convex probes, provide a lower image quality instead, but a greater depth of field. To resolve these differences, assuming measurement independence by zone and patient for this case, we proceed to evaluate two different methodologies (as was mentioned previously) of agreement.

For the descriptive analysis, mean (MD) and standard deviation (SD) or median and interquartile range (IQR), with corresponding 95% confidence interval (CI), were calculated for the quantitative variables. Qualitative variables were described by proportions. Cochran’s Q was applied for comparisons of the probes in each of the 12 quadrants, and for the comparison between the mean LUS of both probes according to quadrants. Student’s t-test (with Levene’s test for equality of variances) was used for variables with normal distribution, and Wilcoxon’s test for variables without normal distribution. The normality of the distributions was verified by the Shapiro-Wilk test. Univariate logistic regression models, receiver operating characteristic (ROC) curves, areas under the curve (AUC), and 95% confidence intervals were calculated to evaluate the predictive performance of both probes for our measured outcomes. Concordance between the results obtained with the convex and the linear probe was studied using Cohen’s Kappa and Delta coefficients for agreement between two raters, determining standard error (SE) and CI. For this, we assume that the regions are independent, which in terms of spatial measurement can be understood as follows. In the analysis all global agreements, marginal agreements, and disagreements are detailed. In addition, approximate confidence intervals have been performed for each estimator, and the overall agreement and positive percentages have been calculated by acquiring each probe. Note that physically the propagation of the ultrasound wave is deeper for the linear case and covers more radius for convex. The significance level was p¡0.05. All analyses were performed with version 24 of the SPSS software package (IBM, Armonk, NY, USA) and Delta Web application and R package deltaMan (5.0) 2022 [34].

### 2.6. Design of the study

The sample size used N, has been introduced from each finding acquisition by region and not by patient. This is due to the aim of discriminating a probe in the study, for each lung ultrasound finding. Thus, we have included a sample size calculation for this research as follows:

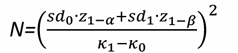

*With* 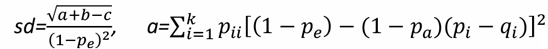 *b=*(1−*p*_*e*_)^2^ ∑*i*≠*j p*_*ij*_ (*q* + *p*_*j*_)^2^, *c=*(*p*_*a*_*p*_*e*_− 2*p*_*e*_+ *p*_*a*_)^2^. Note that when k is equal to 2, p2 = 1−p1, q2 = 1−q1, pe = p1q1 +p2q2, and pa = pe +κ(1−pe), p22 = (pa −p1 +q2)/2, p11 = pa −p22, p12 = p1 −p11, and p21 = q1 −p11. κ0 and κ1 are the contrast values of agreements, and z is the normal distribution value for 1−α and 1−β [35].

We also introduce the kappa sample size estimator with R via the kappaSize library with 0.05 error I and 0.80 of power. With these determinations, we obtain, at least 171 regions of sample size with this analysis [36, 37]. Furthermore, we have also introduced the mean differences between patients, this should be satisfied for considering an effect size of 0.7 and a power of 80% with an error type I of 5%. For this case, the minimum sample required is 16 in a Mann-Whitney non-parametric test methodology. This could be also scalable to the χ2-test in the case of the comparison between categorical variables and 1 degree of freedom. To decrease the effect size, we would have to increase the size of the sample.

Note that the acquisitions have been measured in a COVID-19 pandemic context, and they were obtained to find a sample of patients to explore the impact of these findings on pulmonary illness. The availability was the maximum due to the resources of the medical doctors doing the research in extra time without a sample size design. These calculations have been performed with G⋆power [38].

## 3. Results

From 5th May to 13th July 2021, from the eligible 342 patients, 16 patients with confirmed SARS-CoV-2 RT-PCR were included in the study, which means 32 pulmonary ultrasound examinations were performed and 384 images analyzed. Patient characteristics and clinical were similar in both groups. The median age of the participants was 48 (IQR 23), with a slight male predominance (9, 56.3%) and a mean body mass index of 30.61 (SD 4.73, CI 28.09-33.13). The mean duration of hospitalization was 12.13 days (SD 14.51, CI 4.39-19.86), 7 patients (43.8%) required oxygen therapy during admission, 2 (12.5%) had to be admitted to ICU, 3 (18.8%) had a poor outcome and only 1 (6.3%) patient died at 28 days follow-up.

The results of the linear and convex transducer findings are detailed in Table 1 and Figure 2. On the comparison between the mean LUS of both probes according to quadrants, there was a statistically significant difference between both probes in the right lower lateral (quadrant 4, Q4, p=0.05), right upper posterior (Q5, p=0.011), right lower posterior (Q6, p=0.07), left lower anterior (Q8, p=0.02) and left upper posterior (Q11, p=0.02) quadrants. The mean total LUS with the convex probe was 15.81 (MD 3.69), and the mean total LUS with the linear probe was 10.81 (MD 5.8), with a statistically significant difference between both of them (p=0.001). The most described findings with the convex probe compared to the linear probe were pleural irregularity (142 vs. 111, p=0.005), pleural thickening (134 vs. 91, p=0.002), B-lines ¿ 3 (90 vs. 42, p¡0.01), coalescing B-lines (57 vs. 25, p=0.002) and subpleural consolidation (74 vs. 45, p=0.008); while pleural effusion was more frequent in the linear probe (7 vs. 18, p=0.026). Pulmonary consolidation was more observed with the convex probe but did not reach statistical significance (12 vs. 4, p=0.120).

**Table 1:**
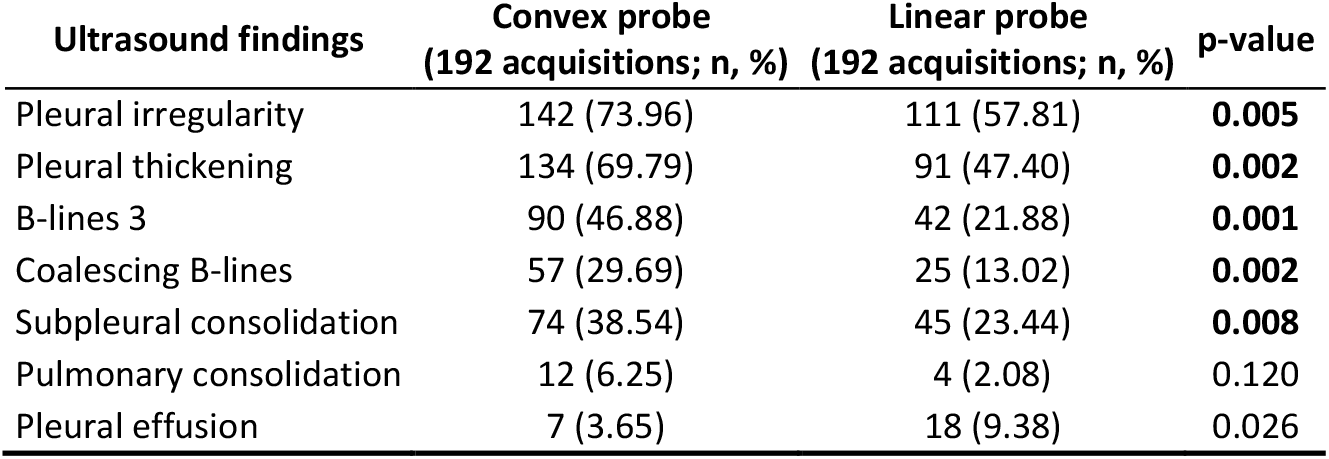
Lung ultrasound findings with convex and linear transducer, comparative with t-student and Wilcoxon’s test between convex and linear transducers.

| Ultrasound findings | Convex probe<br>(192 acquisitions; n, %) | Linear probe<br>(192 acquisitions; n, %) | p-value |
| --- | --- | --- | --- |
| Pleural irregularity | 142 (73.96) | 111 (57.81) | **0.005** |
| Pleural thickening | 134 (69.79) | 91 (47.40) | **0.002** |
| B-lines $\geq 3$ | 90 (46.88) | 42 (21.88) | **0.001** |
| Coalescing B-lines | 57 (29.69) | 25 (13.02) | **0.002** |
| Subpleural consolidation | 74 (38.54) | 45 (23.44) | **0.008** |
| Pulmonary consolidation | 12 (6.25) | 4 (2.08) | 0.120 |
| Pleural effusion | 7 (3.65) | 18 (9.38) | 0.026 |

**Figure 2:**
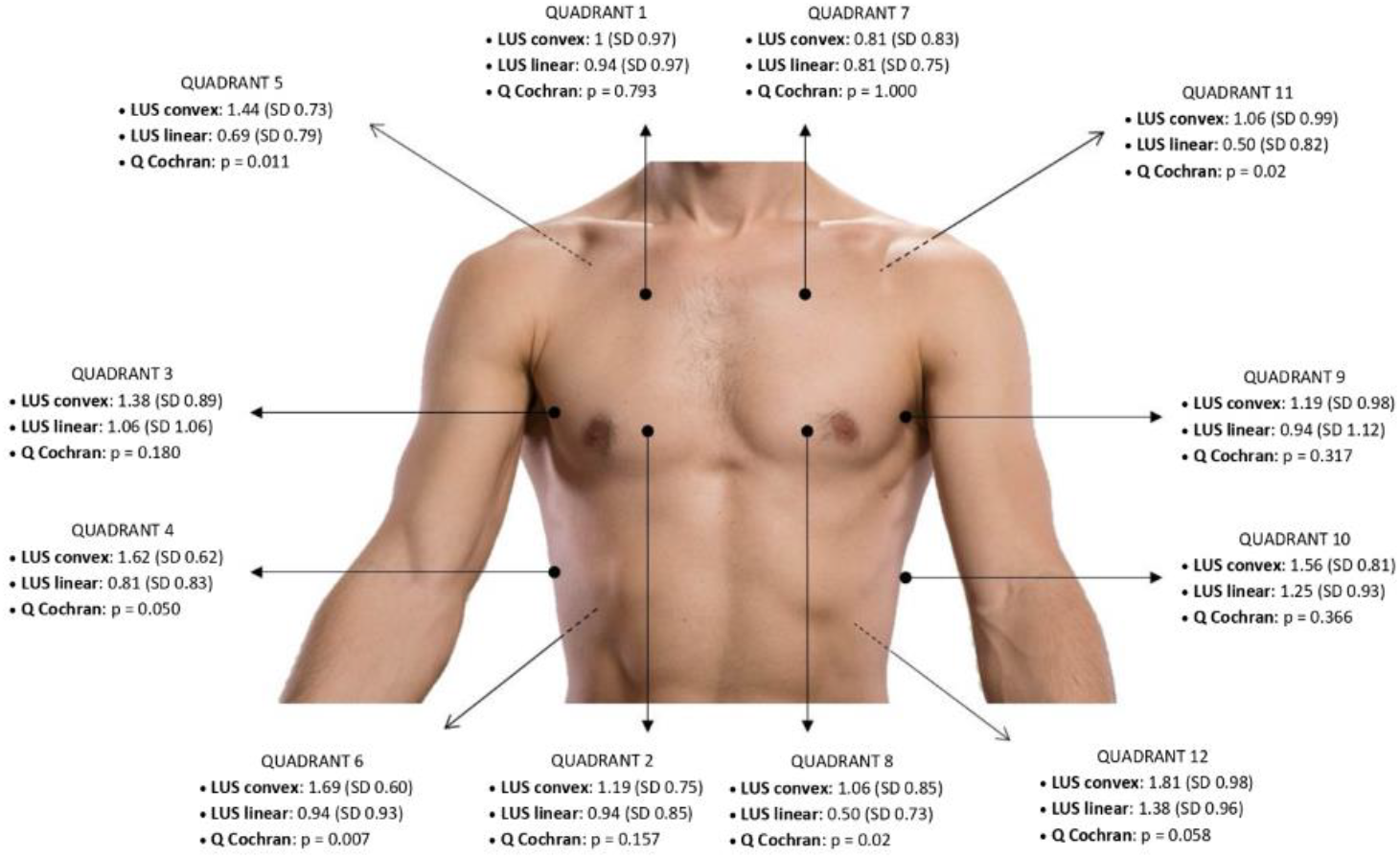
Lung quadrants distribution and comparative between mean LUS convex vs. mean LUS linear by Cochran’s Q test.

Hereafter, we used a global concordance analysis of the lung findings present in the study. We have observed a bad agreement between probes for all the described ultrasound findings in Tables 2–9. Kappa estimator was < 0.4 when we compared LUS (0.04, SE 0.05, CI 0-0.13), pleural irregularity (0.29, SE 0.06, CI 0.16-0.41), pleural thickening (0.25, SE 0.6, CI 0.13-0.37), B-lines > 3 (0.39, SE 0.06, CI 0.27-0.51), coalescing B-lines (0.12, SE 0.07, CI 0-0.26), subpleural consolidation (0.29, SE 0.7, CI 0.16-0.42), pulmonary consolidation (0.10, SE 0.11, CI 0-0.32) and pleural effusion (0.14, SE 0.10, CI 0-0.34). Furthermore, the Delta estimator was < 0.5 when we compared LUS (0.14, SE 0.05, CI 0.04-0.24), pleural irregularity (0.33, SE 0.06, CI 0.20-0.45), pleural thickening (0.28, SE 0.6, CI 0.15-0.40), B-lines > 3 (0.49, SE 0.06, CI 0.38-0.61), coalescing B-lines (0.46, SE 0.06, CI 0.35-0.58) and subpleural consolidation (0.43, SE 0.6, CI 0.31-0.54). However, we observed a Delta score > 0.7 regarding pulmonary consolidation (0.88, SE 0.03, CI 0.81-0.94) and pleural effusion (0.78, SE 0.04, CI 0.70-0.86). This is due to the agreement of negative measurements since there are almost no image phenomena observed in both cases. Delta is detecting this marginal agreement while Kappa it is not considering these quantities. Finally, the percentage of positives for all the linear transducers. This difference was remarkably higher for coalescing B-lines (25.88% positives with the convex probe vs. 12.72% with the linear probe) and pulmonary consolidation (5.26% vs. 1.75% respectively).

**Table 2:** LUS compared with Kappa and Delta estimators and Confidence Intervals at 95% for agreement between convex and linear transducers.

| LUS |  | Linear |  |  |  |  |  |  |
| --- | --- | --- | --- | --- | --- | --- | --- | --- |
|  |  | POS | NEG | TOTAL |  | Punctual Estimation | SE | CI (95%) |
| Convex | POS | 154 | 124 | 278 | Kappa | 0.04 | 0.07 | (0;0.1302) |
|  | NEG | 42 | 42 | 84 | % Concordance | 65.94 |  |  |
|  | TOTAL | 196 | 166 | 362 | Delta | 0.14 | 0.05 | (0.0391;0.2429) |
|  |  |  |  |  | % Linear Positives | 54.14 |  |  |
|  |  |  |  |  | % Convex Positives | 76.80 |  |  |

**Table 3:**
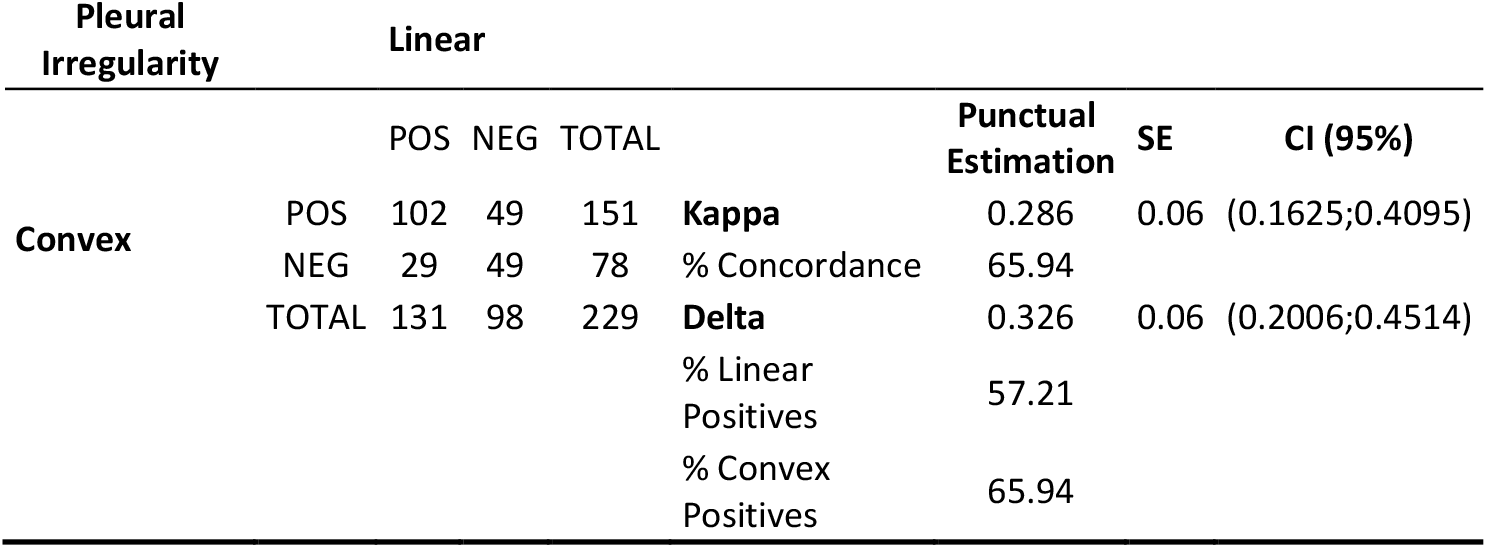
Pleural Irregularity findings compared with Kappa and Delta estimators and Confidence Intervals at 95% for agreement between convex and linear transducers.

**Table 4:** Pleural Thickening findings compared with Kappa and Delta estimators and Confidence Intervals at 95% for agreement between convex and linear transducers.

| Pleural Thickening |  | Linear |  |  |  |  |  |  |
| --- | --- | --- | --- | --- | --- | --- | --- | --- |
|  |  | POS | NEG | TOTAL |  | Punctual Estimation | SE | CI (95%) |
| Convex | POS | 80 | 66 | 146 | **Kappa** | 0.25 | 0.06 | (0.1344;0.3656) |
|  | NEG | 22 | 60 | 82 | % Concordance | 61.40 |  |  |
|  | TOTAL | 102 | 126 | 228 | **Delta** | 0.28 | 0.06 | (0.1515;0.3985) |
|  |  |  |  |  | % Linear Positives | 44.74 |  |  |
|  |  |  |  |  | % Convex Positives | 64.04 |  |  |

**Table 5:** B-lines >3 findings compared by Kappa and Delta estimators and Confidence Intervals at 95% for agreement between convex and linear transducers.

| B-lines<br>>3 |  | Linear |  |  |  | Punctual<br>Estimation | SE | CI (95%) |
| --- | --- | --- | --- | --- | --- | --- | --- | --- |
|  |  | POS | NEG | TOTAL |  |  |  |  |
| Convex | POS | 43 | 50 | 93 | **Kappa** | 0.39 | 0.06 | (0.2734;0.5086) |
|  | NEG | 13 | 123 | 136 | % Concordance | 72.49 |  |  |
|  | TOTAL | 56 | 173 | 229 | **Delta** | 0.49 | 0.06 | (0.3823;0.6057) |
|  |  |  |  |  | % Linear Positives | 24.54 |  |  |
|  |  |  |  |  | % Convex Positives | 40.61 |  |  |

**Table 6:** Coalescing B-lines findings compared by Kappa and Delta estimators and Confidence Intervals at 95% for agreement between convex and linear transducers.

| Coalescing<br>B-lines |  | Linear |  |  |  | Punctual<br>Estimation | SE | CI (95%) |
| --- | --- | --- | --- | --- | --- | --- | --- | --- |
|  |  | POS | NEG | TOTAL |  |  |  |  |
| Convex | POS | 12 | 47 | 59 | **Kappa** | 0.12 | 0.07 | (0;0.2563) |
|  | NEG | 17 | 152 | 169 | % Concordance | 71.93 |  |  |
|  | TOTAL | 29 | 199 | 228 | **Delta** | 0.46 | 0.06 | (0.3503;0.5777) |
|  |  |  |  |  | % Linear Positives | 12.72 |  |  |
|  |  |  |  |  | % Convex Positives | 25.88 |  |  |

**Table 7:** Subpleural Consolidation findings compared by Kappa and Delta estimators and Confidence Intervals at 95% for agreement between convex and linear transducers.

| Subpleural<br>Consolidation |  | Linear |  |  |  | Punctual<br>Estimation | SE | CI (95%) |
| --- | --- | --- | --- | --- | --- | --- | --- | --- |
|  |  | POS | NEG | TOTAL |  |  |  |  |
| Convex | POS | 31 | 45 | 76 | **Kappa** | 0.29 | 0.07 | (0.1557;0.4183) |
|  | NEG | 22 | 132 | 154 | % Concordance | 70.87 |  |  |
|  | TOTAL | 53 | 177 | 230 | **Delta** | 0.429 | 0.06 | (0.3134;0.5446) |
|  |  |  |  |  | % Linear<br>Positives | 23.04 |  |  |
|  |  |  |  |  | % Convex<br>Positives | 33.04 |  |  |

**Table 8:**
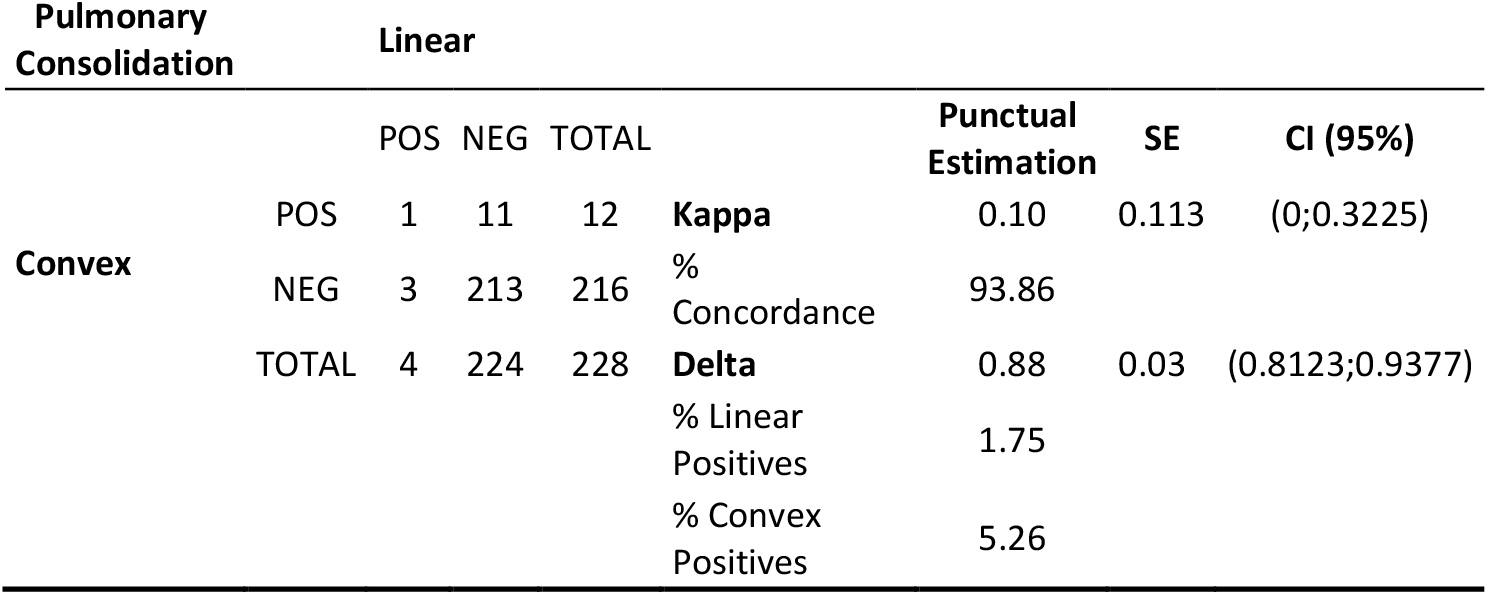
Pulmonary Consolidation findings compared by Kappa and Delta estimators and Confidence Intervals at 95% for agreement between convex and linear transducers.

**Table 9:**
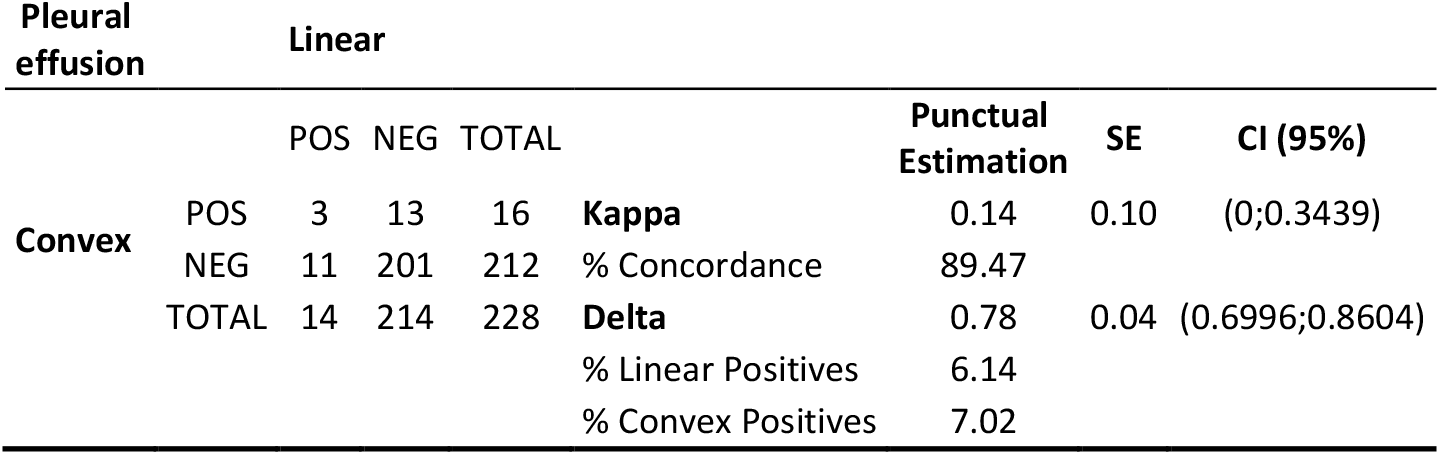
Pleural Effusion consolidation findings compared by Kappa and Delta estimators and Confidence Intervals at 95% for agreement between convex and linear transducers.

The data obtained revealed a low correlation in the description of the studied findings between the two selected probes. This difference is less pronounced in the description of pleural irregularity and pleural effusion, but even so, these still showed Kappa values <0.4 and Delta values <0.5 in the majority of the cases and 0.79 and 0.87 for the cases where there are almost no indices of Pulmonary Consolidation or Pleural Effusion.

The analysis of the measured outcomes with both probes showed that the odds ratios, ROC curves, and AUC demonstrated a positive trend in the prediction of these events. However, the low number of events measured resulted in CIs that were too wide to be considered clinically or statistically significant.

## 4. Discussion

The published literature about lung ultrasound in COVID-19 is constantly increasing. However, the choice of the transducer is an unresolved issue [18]. The two most studied probes are the low-frequency convex and the high-frequency linear probe [5]. Lung ultrasound aims to detect artifacts generated by pneumonia over the lung parenchyma. The image received by the transducers will differ according to the frequency at which these ultrasounds are emitted. High-frequency probes (like the linear transducer) provide higher resolution, but a shallower depth of field. Low-frequency transducers (like the convex probe) provide a lower image quality instead, but a greater depth of field [28].

Soldati’s proposal for lung ultrasound standardization in COVID-19 remains as the reference document [39], but it only indicates the choice between convex and linear transducers according to body size [20]. Several studies agree on the choice of the linear probe for assessing the presence of pleural affections or evaluating children and thin patients; while the convex probe is reserved for the study of the pulmonary parenchyma and the remaining patients [39, 27]. Other articles just suggest the convex probe without explaining why it should be used [40]. Outside COVID-19, the use of one or the other probe also generates discussion [41]. Both probes are therefore suitable and useful for the assessment of COVID-19 pneumonia, but there are no comparative studies to justify these indications.

The latest international guidelines reiterate the recommendation for the choice of a transducer according to body phenotype, being a convex transducer preferable for subpleural parenchyma and a linear transducer for pleural alterations [26]. For this recommendation, however, they rely on a 2013 article that does not refer to this issue [42], since to our knowledge there are no such studies. Furthermore, the aforementioned international guideline about subpleural consolidations equates both probes, without any study or theoretical justification to support this recommendation [26]. This contradicts previous studies that have already demonstrated, in the case of B lines, significant differences between both transducers [43, 44]. These articles conclude that lung artifacts detected on lung ultrasound are highly dependent on the transducer used and these are not interchangeable.

Based on our findings, we corroborate that lung ultrasound in COVID-19 is a transducer-dependent technique, with significant differences between the findings of one probe and the other [45, 46]. Therefore, the results of each transducer are not comparable, and LUS cut-off points for disease severity should be adjusted appropriately. This difference is most pronounced in the posterior quadrants and the cardiac area, probably due to the lower depth of field of the linear probe to overcome the accumulation of adipose tissue and the obstacle of the heart. It is noteworthy that the convex probe shows superiority in the detection of pleural pathology (pleural irregularity, pleural thickening, and subpleural consolidation) excluding pleural effusion. It is also significantly superior in the detection of ¿3 B-lines and lung consolidation as postulated, although the last one did not reach statistical significance. These findings show that the convex probe obtained a greater number of findings and a higher LUS, which probably indicates that it is more sensitive. In contrast, we found a better performance of the linear transducer in the detection of pleural effusion. Note that the Delta estimator is quite high in Table 8 and Table 9. This is due to the agreement of negative measurements, there are almost no image phenomena observed in this case around 12, and Delta is detecting this marginal negative agreement. Then, the acquisition shows almost no pulmonary consolidations, and even in this case, the convex probe detects more positives.

This is the first research comparing COVID-19 lung ultrasound findings in the same patient and at the same time with two different transducers. Moreover, we performed a strict protocol to reduce the inter-operator variability in image acquisition, the possible preference or ability of the operator for one or the other probe, the obtainment of images at different stages, and the influence of other analytical or radiological severity data on the acquisition of images. It has been proven in several studies that inter-rater reliability is poor in the assessment of specific lung ultrasound findings in COVID-19 patients [47, 48, 49]. However, the use of ultrasound assessment scores seems to reduce this problem [50, 48] but only with specific training in COVID-19 pneumonia [47].

Besides, in our case, for each ultrasound finding, we have at least 228 regions to analyse. So, the sample size seems to be enough to corroborate the conclusion. In summary, no comparative studies between linear and convex probes have been reported to date, at least in the field of lung ultrasound in viral pneumonia-like COVID-19. For these reasons, we believe that this article may be of interest to the scientific community by supporting some of its recommendations with objective data. Likewise, it will also generate new hypotheses on some of the current guidelines that are considered to be immovable.

Among the limitations of this work, the strict acquisition protocol led to an increased time spent with each patient, therefore reducing the size of the sample. Likewise, the low number of patients included implied that the measured events were so infrequent that no conclusions could be reached about the association with our outcomes (poor outcome or 28-day mortality). Therefore, we cannot conclude that the use of one transducer over another one is associated with better clinical outcomes. On the other hand, the relative youth of our patients may have resulted in lower mortality, making it difficult to find differences in these outcomes. Additionally, the absence of another reference imaging test (e.g., CT) makes it difficult to state categorically that one probe is more sensitive than the other. Finally, although the coexistence of pulmonary superinfection was an exclusion criterion, the absence of pathognomonic data (B-line, subpleural consolidations, and pleural effusion) confer to these signs a variable specificity [7]. Differences in lung involvement of new variants (e.g., omicron) should be studied in future investigations.

## 5. Conclusions

To conclude, lung ultrasound in COVID-19 is a transducer-dependent technique, with poor concordance in the description of ultrasound findings between convex and linear transducers. This difference is more pronounced in the posterior and cardiac areas. The convex probe is superior to the linear probe in detecting pleural pathology, excluding pleural effusion, and in the detection of > 3 B-lines or lung consolidation. Although both linear and convex transducers are suitable for evaluation for COVID-19 pneumonia, the convex transducer provides more information. Larger studies are required to better confirm an expected higher sensitivity of the convex probe and to demonstrate a different predictive value between both transducers.

## Data Availability

All data produced in the present study are available upon reasonable request to the authors

## Acknowledgments

To Valme Sánchez Cabrera, for her support in managing the database, to Alejandra Ciria García, for her assistance in the text edition, to Guillermo Rus for sharing his knowledge of ultrasonic probes, Juan de Dios Luna and Pedro Femia for their statistical advice, and to all the healthcare professionals involved in the COVID-19 assistance.

## References

[1] COVID-19 Map. Johns Hopkins Coronavirus Resource Center n.d. https://coronavirus.jhu.edu/map.html (accessed October 10, 2021).

[2] Mehta OP, Bhandari P, Raut A, Kacimi SEO, Huy NT. Coronavirus disease (COVID-19): comprehensive review of clinical presentation. Front Public Health 2021;8:582932. 10.3389/fpubh.2020.582932.

[3] Landini N, Orlandi M, Fusaro M, Ciet P, Nardi C, Bertolo S, et al. The Role of Imaging in COVID-19 Pneumonia Diagnosis and Management: Main Positions of the Experts, Key Imaging Features and Open Answers. J Cardiovasc Echogr 2020;30:S25–30. 10.4103/jcecho.jcecho_59_20.

[4] Nouvenne A, Zani MD, Milanese G, Parise A, Baciarello M, Bignami EG, et al. Lung Ultrasound in COVID-19 Pneumonia: Correlations with Chest CT on Hospital admission. Respiration 2020;99:617–24. 10.1159/000509223.

[5] Peixoto AO, Costa RM, Uzun R, Fraga AMA, Ribeiro JD, Marson FAL. Applicability of lung ultrasound in COVID-19 diagnosis and evaluation of the disease progression: A systematic review. Pulmonology 2021;27:529–62. 10.1016/j.pulmoe.2021.02.004.

[6] Guzmán-García MB, Mohedano-Moriano A, González-González J, Morales-Cano JM, Campo-Linares R, Lozano-Suárez C, et al. Lung Ultrasound as a Triage Method in Primary Care for Patients with Suspected SARS-CoV-2 Pneumonia. JCM 2022;11:6420. 10.3390/jcm11216420.

[7] Allinovi M, Parise A, Giacalone M, Amerio A, Delsante M, Odone A, et al. Lung Ultrasound May Support Diagnosis and Monitoring of COVID-19 Pneumonia. Ultrasound in Medicine & Biology 2020;46:2908–17. 10.1016/j.ultrasmedbio.2020.07.018.

[8] Gil-Rodríguez J, Martos-Ruiz M, Benavente-Fernández A, Aranda-Laserna P, Montero-Alonso MÁ, Peregrina-Rivas J-A, et al. Lung ultrasound score severity cut-off points in COVID-19 pneumonia. A systematic review and validating cohort. Medicina Clínica 2023;160:531–9. 10.1016/j.medcli.2023.01.024.

[9] Ianniello S, Piccolo CL, Buquicchio GL, Trinci M, Miele V. First-line diagnosis of paediatric pneumonia in emergency: lung ultrasound (LUS) in addition to chest-X-ray (CXR) and its role in follow-up. BJR 2016;89:20150998. 10.1259/bjr.20150998.

[10] Wang Y, Zhang Y, He Q, Liao H, Luo J. Quantitative Analysis of Pleural Line and B-Lines in Lung Ultrasound Images for Severity Assessment of COVID-19 Pneumonia. IEEE Trans Ultrason, Ferroelect, Freq Contr 2022;69:73–83. 10.1109/TUFFC.2021.3107598.

[11] Kameda T, Mizuma Y, Taniguchi H, Fujita M, Taniguchi N. Point-of-care lung ultrasound for the assessment of pneumonia: a narrative review in the COVID-19 era. J Med Ultrasonics 2021;48:31–43. 10.1007/s10396-020-01074-y.

[12] Lomoro P, Verde F, Zerboni F, Simonetti I, Borghi C, Fachinetti C, et al. COVID-19 pneumonia manifestations at the admission on chest ultrasound, radiographs, and CT: single-center study and comprehensive radiologic literature review. European Journal of Radiology Open 2020;7:100231. 10.1016/j.ejro.2020.100231.

[13] Chinese Critical Care Ultrasound Study Group (CCUSG), Peng Q-Y, Wang X-T, Zhang L-N. Findings of lung ultrasonography of novel corona virus pneumonia during the 2019–2020 epidemic. Intensive Care Med 2020;46:849–50. 10.1007/s00134-020-05996-6.

[14] Mazzola M, Pugliese NR, Zavagli M, De Biase N, Bandini G, Barbarisi G, et al. Diagnostic and Prognostic Value of Lung Ultrasound B-Lines in Acute Heart Failure With Concomitant Pneumonia. Front Cardiovasc Med 2021;8:693912. 10.3389/fcvm.2021.693912.

[15] Caiulo VA, Gargani L, Caiulo S, Fisicaro A, Moramarco F, Latini G, et al. Lung ultrasound characteristics of community-acquired pneumonia in hospitalized children. Pediatric Pulmonology 2013;48:280–7. 10.1002/ppul.22585.

[16] Cho Y-J, Song K-H, Lee Y, Yoon JH, Park JY, Jung J, et al. Lung ultrasound for early diagnosis and severity assessment of pneumonia in patients with coronavirus disease 2019. Korean J Intern Med 2020;35:771–81. 10.3904/kjim.2020.180.

[17] Boero E, Rovida S, Schreiber A, Berchialla P, Charrier L, Cravino MM, et al. The COVID-19 Worsening Score (COWS)—a predictive bedside tool for critical illness. Echocardiography 2021;38:207–16. 10.1111/echo.14962.

[18] Gil-Rodríguez J, Martos-Ruiz M, Peregrina-Rivas J-A, Aranda-Laserna P, Benavente-Fernández A, Melchor J, et al. Lung Ultrasound, Clinical and Analytic Scoring Systems as Prognostic Tools in SARS-CoV-2 Pneumonia: A Validating Cohort. Diagnostics 2021;11:2211. 10.3390/diagnostics11122211.

[19] Carrer L, Donini E, Marinelli D, Zanetti M, Mento F, Torri E, et al. Automatic Pleural Line Extraction and COVID-19 Scoring From Lung Ultrasound Data. IEEE Trans Ultrason, Ferroelect, Freq Contr 2020;67:2207–17. 10.1109/TUFFC.2020.3005512.

[20] Soldati G, Smargiassi A, Inchingolo R, Buonsenso D, Perrone T, Briganti DF, et al. Proposal for International Standardization of the Use of Lung Ultrasound for Patients With COVID-19: A Simple, Quantitative, Reproducible Method. J of Ultrasound Medicine 2020;39:1413–9. 10.1002/jum.15285.

[21] Rouby J-J, Arbelot C, Gao Y, Zhang M, Lv J, An Y, et al. Training for Lung Ultrasound Score Measurement in Critically Ill Patients. Am J Respir Crit Care Med 2018;198:398–401. 10.1164/rccm.201802-0227LE.

[22] Soummer A, Perbet S, Brisson H, Arbelot C, Constantin J-M, Lu Q, et al. Ultrasound assessment of lung aeration loss during a successful weaning trial predicts postextubation distress*: Critical Care Medicine 2012;40:2064–72. 10.1097/CCM.0b013e31824e68ae.

[23] Blazic I, Cogliati C, Flor N, Frija G, Kawooya M, Umbrello M, et al. The use of lung ultrasound in COVID-19. ERJ Open Res 2023;9:00196–2022. 10.1183/23120541.00196-2022.

[24] Song G, Qiao W, Wang X, Yu X. Association of Lung Ultrasound Score with Mortality and Severity of COVID-19: A Meta-Analysis and Trial Sequential Analysis. International Journal of Infectious Diseases 2021;108:603–9. 10.1016/j.ijid.2021.06.026.

[25] Sun Z, Zhang Z, Liu J, Song Y, Qiao S, Duan Y, et al. Lung Ultrasound Score as a Predictor of Mortality in Patients With COVID-19. Front Cardiovasc Med 2021;8:633539. 10.3389/fcvm.2021.633539.

[26] Demi L, Wolfram F, Klersy C, De Silvestri A, Ferretti VV, Muller M, et al. New International Guidelines and Consensus on the Use of Lung Ultrasound. J of Ultrasound Medicine 2023;42:309–44. 10.1002/jum.16088.

[27] European Society of Radiology (ESR), Clevert D-A, Sidhu PS, Lim A, Ewertsen C, Mitkov V, et al. The role of lung ultrasound in COVID-19 disease. Insights Imaging 2021;12:81. 10.1186/s13244-021-01013-6.

[28] Bobbia X, Chabannon M, Chevallier T, De La Coussaye JE, Lefrant JY, Pujol S, et al. Assessment of five different probes for lung ultrasound in critically ill patients: A pilot study. The American Journal of Emergency Medicine 2018;36:1265–9. 10.1016/j.ajem.2018.03.077.

[29] Buda N, Skoczylas A, Demi M, Wojteczek A, Cylwik J, Soldati G. Clinical Impact of Vertical Artifacts Changing with Frequency in Lung Ultrasound. Diagnostics 2021;11:401. 10.3390/diagnostics11030401.

[30] Szabo TL, Lewin PA. Ultrasound Transducer Selection in Clinical Imaging Practice. Journal of Ultrasound in Medicine 2013;32:573–82. 10.7863/jum.2013.32.4.573.

[31] Liu J, Guo G, Kurepa D, Volpicelli G, Sorantin E, Lovrenski J, et al. Specification and guideline for technical aspects and scanning parameter settings of neonatal lung ultrasound examination. The Journal of Maternal-Fetal & Neonatal Medicine 2022;35:1003–16. 10.1080/14767058.2021.1940943.

[32] Collins GS, Reitsma JB, Altman DG, Moons KGM. Transparent Reporting of a multivariable prediction model for Individual Prognosis Or Diagnosis (TRIPOD): The TRIPOD Statement. Ann Intern Med 2015;162:55–63. 10.7326/M14-0697.

[33] Mento F, Perrone T, Macioce VN, Tursi F, Buonsenso D, Torri E, et al. On the Impact of Different Lung Ultrasound Imaging Protocols in the Evaluation of Patients Affected by Coronavirus Disease 2019: How Many Acquisitions Are Needed? J Ultrasound Med 2021;40:2235–8. 10.1002/jum.15580.

[34] Andrés AM, Marzo PF. Delta: A new measure of agreement between two raters. British Journal of Mathematical and Statistical Psychology 2004;57:1–19. 10.1348/000711004849268.

[35] Bujang MA, Baharum N. Guidelines of the minimum sample size requirements for Kappa agreement test. Ebph 2022;14. 10.2427/12267.

[36] Rotondi MA. kappaSize: Sample Size Estimation Functions for Studies of Interobserver Agreement 2018.

[37] Rotondi MA, Donner A. A confidence interval approach to sample size estimation for interobserver agreement studies with multiple raters and outcomes. Journal of Clinical Epidemiology 2012;65:778–84. 10.1016/j.jclinepi.2011.10.019.

[38] Kang H. Sample size determination and power analysis using the G*Power software. J Educ Eval Health Prof 2021;18:17. 10.3352/jeehp.2021.18.17.

[39] Buda N, Andruszkiewicz P, Czuczwar M, Gola W, Kosiak W, Nowakowski P, et al. Consensus of the Study Group for Point-of-Care Lung Ultrasound in the intensive care management of COVID-19 patients. Ait 2020;52:83–90. 10.5114/ait.2020.96560.

[40] Gargani L, Soliman-Aboumarie H, Volpicelli G, Corradi F, Pastore MC, Cameli M. Why, when, and how to use lung ultrasound during the COVID-19 pandemic: enthusiasm and caution. European Heart Journal - Cardiovascular Imaging 2020;21:941–8. 10.1093/ehjci/jeaa163.

[41] Heldeweg MLA, Lopez Matta JE, Haaksma ME, Smit JM, Elzo Kraemer CV, De Grooth H-J, et al. Lung ultrasound and computed tomography to monitor COVID-19 pneumonia in critically ill patients: a two-center prospective cohort study. ICMx 2021;9:1. 10.1186/s40635-020-00367-3.

[42] Smargiassi A, Inchingolo R, Soldati G, Copetti R, Marchetti G, Zanforlin A, et al. The role of chest ultrasonography in the management of respiratory diseases: document II. Multidis Res Med 2013;8. 10.4081/mrm.2013.556.

[43] Sperandeo M, Varriale A, Sperandeo G, Polverino E, Feragalli B, Piattelli ML, et al. Assessment of ultrasound acoustic artifacts in patients with acute dyspnea: a multicenter study. Acta Radiol 2012;53:885–92. 10.1258/ar.2012.120340.

[44] Dietrich CF, Mathis G, Blaivas M, Volpicelli G, Seibel A, Wastl D, et al. Lung B-line artefacts and their use. J Thorac Dis 2016;8:1356–65. 10.21037/jtd.2016.04.55.

[45] Altman DG. Practical statistics for medical research. Boca Raton, Fla: Chapman & Hall/CRC; 1999.

[46] Andrés AM, Marzo PF. Delta: A new measure of agreement between two raters. British Journal of Mathematical and Statistical Psychology 2004;57:1–19. 10.1348/000711004849268.

[47] Šustic A, Miroševic M, Szuldrzynski K, Marčun R, Haznadar M, Podbegar M, et al. Inter-observer reliability for different point-of-care lung ultrasound findings in mechanically ventilated critically ill COVID-19 patients. J Clin Monit Comput 2022;36:279–81. 10.1007/s10877-021-00726-9.

[48] Herraiz JL, Freijo C, Camacho J, Muñoz M, González R, Alonso-Roca R, et al. Inter-Rater Variability in the Evaluation of Lung Ultrasound in Videos Acquired from COVID-19 Patients. Applied Sciences 2023;13:1321. 10.3390/app13031321.

[49] Baloescu C, Chen A, Schnittke N, Hicks B, Zhu M, Kaili M, et al. Development and interobserver reliability of a rating scale for lung ultrasound pathology in lower respiratory tract infection. WFUMB Ultrasound Open 2023;1:100006. 10.1016/j.wfumbo.2023.100006.

[50] Li W, Fu M, Qian C, Liu X, Zeng L, Zhou H, et al. Bedside lung ultrasound score (LUSS) on assessing pneumonia in COVID-19 neonates. Preprints; 2020. 10.22541/au.160091445.58861962.

